# Prevalence of female athlete triad (FAT) in rhythmic gymnastics, an aesthetic sport, and its association with risks of body-image distortion and eating disorders

**DOI:** 10.1101/2024.10.29.24316350

**Authors:** Kayo Yoshitani-Kuwabara, Yukina Yumen, Yumi Takayama, Natsuho Kitayama, Fumiaki Hanzawa, Naoki Sakane, Narumi Nagai

## Abstract

**Background:** Female athlete triad (FAT), a syndrome linking low energy availability (LEA), menstrual dysfunction (MD), and impaired bone health, has serious potential consequences for sports performance and health. In this cross-sectional study, we investigated prevalences of each FAT component in female athletes of aesthetic and non-aesthetic sports and in non-athlete females, and its association with body image (BI) distortion and eating behaviors.

**Methods:** Google Forms was used to examine self-reported height and weight, menstruation conditions, history of stress fractures (HSF), BI, the Eating Attitude Test (EAT)-26 (indicator of eating disorder) scores, and eating behaviors; these attributes were compared between 3 groups (athletes of aesthetic sports [rhythmic gymnasts, *n=*40] and non-esthetic sports [volleyball players, *n=*46]; other females of the same age group [non-athletes, *n=*108]). To identify risk factors for each FAT component, multivariate logistic-regression analyses were used. LEA, MD, and HSF were dependent variables; rhythmic gymnasts, distorted BI, and EAT-26 scores were independent variables.

**Results:** Rhythmic gymnasts had a lower body mass index (*p<*0.001), higher LEA and MD prevalence (both *p<*0.001), and greater percentage of participants with ≥2 FAT criteria (*p<*0.05) compared with other groups; exhibited higher distorted BI scores (*p<*0.05) than volleyball players; and tended to overestimate their body shape. Regarding eating behaviors, rhythmic gymnasts had higher EAT-26 scores (*p<*0.001) than other groups and significantly tended to skip breakfast (*p<*0.05) than non-athletes. Multivariate logistic-regression analysis revealed that rhythmic gymnasts and body-shape overestimation were independently associated with LEA; rhythmic gymnasts and high EAT-26 scores were independently associated with MD.

**Conclusions:** FAT prevalence was approximately 1.5 times higher among rhythmic gymnasts than among volleyball players and non-athletes. These findings suggest that rhythmic gymnasts are at a higher risk of FAT and being an athlete in aesthetic sports, body-shape overestimation, and higher EAT-26 scores are risk factors for FAT.

## Introduction

An increasing number of women are engaging in sports from a young age [1]. However, long-term rigorous training and weight loss to enhance athletic performance may predispose young women to the risk of developing female athlete triad (FAT) [2,3]. This term encompasses a range of health-related issues relating to low energy availability (LEA) resulting from an energy deficit when reaching a certain threshold level, leading to menstrual dysfunction (MD) and low bone mineral density as a result of reduced bone formation associated with menstrual irregularities during adolescence [2]. LEA is characterized as increased energy expenditure owing to intense training alongside an insufficient energy intake [3].

The prevalence of FAT varies with sport characteristics and is notably high in aesthetic sports such as rhythmic gymnastics and figure skating [2]. We therefore focused on female rhythmic gymnasts, a group with approximately 10,000 participants in Japan, who often continue their competitive careers from elementary school through to high school and university during their growth period [4]. One study involving Japanese female elite athletes [5] reported that the rate of amenorrhea among female rhythmic gymnasts was significantly higher at 40.9%, when compared with 6.2% and 3.8% in volleyball and soccer players, respectively. Moreover, the prevalence of stress fractures was higher among rhythmic gymnasts at 45.5%, when compared with 16.0% and 7.5% in volleyball and soccer players, respectively.

Rhythmic gymnastics is a sport where being physically thin has both competitive and aesthetic value. Athletes in this discipline have been known to often restrict their diets to enhance performance [6], leading to a higher prevalence of LEA and resulting in amenorrhea and stress fractures. To protect these athletes, it is crucial to recognize and address FAT-related signs early. In previous research, we showed that the desire to lose weight is linked to a distorted body image (BI) perception among Japanese non-athlete women [7]. This distorted perception may also cause rhythmic gymnasts to pursue excessive thinness, worsening the issues in relation to FAT. Investigating the association between a desire to lose weight and BI in rhythmic gymnasts could help address this issue. To better understand BI and the eating behaviors of rhythmic gymnasts, we compared such gymnasts with female athletes of non-aesthetic sports and with non-athletes. The most appropriate age group for this comparison involves female high school and university students, as their menstrual cycles are stabilized and their growth and development are nearing completion.

This study aimed to elucidate the characteristics and prevalence of FAT and BI, eating behaviors, and tendencies toward eating disorder in female rhythmic gymnasts in high school or at university in comparison with female volleyball players and non-athletes of similar age. We investigated the prevalence of each FAT component female athletes of aesthetic and non-esthetic sports and non-athlete females, and its association with distortion and eating behaviors.

## Methods

### Study design and population

This cross-sectional study was conducted using a web-based survey. Participants included female high school and university students engaged in aesthetic sports (rhythmic gymnasts, *n*=40), non-aesthetic sports (volleyball players, *n*=46), and non-athlete female students of the same age (*n*=108). Rhythmic gymnasts were recruited between 15 April and 1 June 2022, while female volleyball players and non-athletes were recruited between 15 June and 16 July 2023 through flyers requesting participation.

The rhythmic gymnasts and volleyball players participating in this study performed at a highly competitive level, capable of competing in prefectural and national tournaments, practicing their sports almost daily, and engaging in high-intensity exercise. Inclusion criteria comprised women who belonged to high-school or university rhythmic gymnastics or volleyball club, who agreed to participate in the study, and who could complete the questionnaire in Japanese.

Non-athlete women included high school students who visited our university’s open campus and university students from our university and other nearby universities in Hyogo Prefecture or surrounding prefectures. Exclusion criteria were men, university or vocational school students majoring in nutrition, and non-athlete women with exercise habits. The study’s purpose, its anonymity, the voluntary nature of participation, and the condition that completing the web survey would be considered consent to participate in the study were stated in the flyers and at the beginning of the web survey. This information was also explained verbally when distributing the flyers. Written informed consent was obtained from the study participants. Data from all respondents who completed the web survey were used for analysis after confirming there were no errors, resulting in a 100% valid response rate.

This study was approved by the Research Ethics Committee of the University of Hyogo (No. 275, March 10, 2022, No. 314, June 7, 2023). The study procedures were conducted in accordance with the Declaration of Helsinki.

### Questionnaire

We developed a self-report questionnaire using Google Forms. Study participants accessed the questionnaire via a flyer with a QR code link. The survey items sought data in relation to basic characteristics, The prevalence of each FAT component, BI, and eating behaviors.

### Basic characteristics

Participants self-reported their school category, age (years), athletic experience (years), height (cm), and weight (kg). Body mass index (BMI) (kg/m^2^) was calculated as weight (kg) divided by height squared (m^2^).

### FAT

LEA was screened using the definition of the American College of Sports Medicine [8], which considers clear signs of low energy status to be a body weight <85% of normal (BMI=22 kg/m^2^).

Menstrual abnormalities were determined according to Obstetrics and Gynecology Clinical Practice Guidelines, Outpatient Gynecology Edition, 2020 [9]. Participants reported their current menstrual status either as: (i) a regular menstrual cycle, (ii) oligomenorrhea (cycle of ≥39 days), (iii) frequent menstruation (cycle of ≤24 days), (iv) amenorrhea (absence of menstruation ≥3 months), (v) irregular, or (vi) no menarche.

Responses ranging from (ii) to (vi) were categorized as menstrual abnormalities. Participants who responded ‘yes’ to having a past experience of a stress fracture (response options: yes/no/don’t know) were considered to have had an HSF.

### BI-related indicators

BI was assessed using the Japanese version of the body image scale (J-BIS) developed and examined for reliability and validity [10], which comprises 10 female silhouettes. Participants selected the silhouette they considered best represented their current-BI (C-BI) and their ideal-BI (I-BI). Using the BMI range (mean±standard error) of each silhouette from a previous study [10], the silhouette number corresponding to a participant’s BMI was defined as the actual-BI (A-BI). The difference between the C-BI and A-BI was defined as ‘BI distortion’ (BI distortion: overestimation if >0, underestimation if <0) [11,12].

The survey also included items on weight control behavior based on BI, such as the frequency of weighing (≥5 times per week, 3–4 times per week, 1–2 times per week, or never) and weight loss experience (currently attempting to lose weight, not currently but have attempted to lose weight in the past, or neither currently nor in the past).

### Eating behaviors

EDs were assessed using the eating attitudes test (EAT-26), a validated screening tool [13,14] for abnormal eating behaviors [15,16]. This test consists of 26 questions rated on a six-point Likert scale ranging from 0 to 3 (3, always; 2, almost always; 1, frequently; 0, rarely, almost never, or never). Results were analyzed using reverse scoring, with a global score of >20 indicating high risk of an ED.

Additional items included self-evaluation of food intake (response options: consistent with consumption, more than consumption, less than consumption, and don’t know), the frequency of skipping breakfast ≥3 times a week (yes/no), and eating dinner ≥2 h before bedtime ≥3 times a week (yes/no). Respondents also reported the frequency of staple food intake, specifying how many times per week they consumed rice (1 bowl per serving), bread (1 slice of a 6-slice loaf per serving), and noodles (1 bowl or 1 plate per serving).

### Sample size

The sample size was calculated using the “pwr” R package, as no previous studies were available to determine the effect size. Thus, the following parameters were used: number of groups = 3, alpha = 0.05, power = 0.8, and effect size = 0.4 (considered large). Based on these inputs, 22 participants per group were required, resulting in a total of 66 participants.

### Statistical analysis

All analyses were conducted using IBM SPSS Statistics version 28.0 for Microsoft Windows (IBM, Tokyo) software. Normality was assessed using a Shapiro-Wilk test prior to performing statistical analyses. Group comparisons were undertaken using one-way analysis of variance (ANOVA) and a Kruskal-Wallis test, with Bonferroni correction applied in the test thereafter. Chi-square tests were used to examine differences between categorical variables. In logistic regression analysis, the dependent variable was FAT (LEA, MD, and HSF; applicable=1, not applicable=0). The explanatory variables included aesthetics (rhythmic gymnastics), which showed the highest FAT prevalence in the three-group comparison, EDs (previously reported to be associated with FAT), and BI distortion (a key research objective). Non-collinearity of the explanatory variables was confirmed using Spearman’s correlation coefficients (all <0.3) and collinearity statistics (variance inflation factor, <10). For comparisons between rhythmic gymnasts and volleyball groups, age and athletic experience were adjusted as potential confounding factors. For comparisons between rhythmic gymnasts and non-athlete groups, only age was adjusted. The significance level was set at <5% (two-sided test).

## Results

### Participant characteristics

Table 1 summarizes the basic characteristics of the female rhythmic gymnasts, volleyball players, and non-athletes. No significant differences were observed in school type, age, or athletic history among the groups. The rhythmic gymnasts were significantly shorter than the volleyball players (*p*<0.05) and had a notably lower mean weight (rhythmic gymnasts [46.3 ± 4.6 kg] vs. volleyball players [60.8±5.9 kg] vs. non-athletes [49.8±6.7 kg]), and BMI (rhythmic gymnasts [18.7±1.7 kg/m²] vs. volleyball players [21.0±1.4 kg/m²] vs. non-athletes [19.9±2.4 kg/m²]) (*p<*0.001).

**Table 1.** Participant characteristics.

|  | Rhythmic<br>gymnasts<br>(*n*=40) | Volleyball<br>players<br>(*n*=46) | Non-Athletes<br>(*n*=108) | *p*-value |
| --- | --- | --- | --- | --- |
| School category |  |  |  | — |
| High school | 23 | 23 | 47 |  |
| University / College | 17 | 23 | 61 |  |
| Age (years) | 17.5± 2.0 | 17.9±2.0 | 17.8±1.6 | 0.488<sup>†</sup> |
| Athletic career (years) | 11.2±3.0 | 10.4±2.7 | — | 0.206<sup>‡</sup> |
| Height (cm) | 157.3±4.6 | 170.0±6.8<sup>a</sup> | 158.0±5.8<sup>b</sup> | < 0.001<sup>Φ</sup> |
| Weight (kg) | 46.3±4.6 | 60.8±5.9<sup>a</sup> | 49.8±6.7<sup>a,b</sup> | < 0.001<sup>Φ</sup> |
| Body Mass Index (kg/m<sup>2</sup>) | 18.7±1.7 | 21.0±1.4<sup>a</sup> | 19.9±2.4<sup>a,b</sup> | < 0.001<sup>Φ</sup> |

Values represent mean±SD.
<sup>†</sup> Kruskal-Wallis. <sup>‡</sup> Paired t-test. <sup>Φ</sup> One-way ANOVA.
<sup>a</sup> *p*<0.05 vs Rhythmic gymnasts, <sup>b</sup> *p*<0.05 vs Volleyball players, by Bonferroni post hoc test in each group.
Height and weight were self-reported.

### Prevalence of FAT

Fig 1A shows the results of a comparison in the prevalence of FAT among the three female groups. The prevalence of LEA (rhythmic gymnasts [51.3%] vs. volleyball players [2.2%] vs. non-athletes [29.0%]) and MD (rhythmic gymnasts [64.1%] vs. volleyball players [43.5%] vs. non-athletes [26.2%]) was significantly higher in the rhythmic gymnasts (*p*<0.001). There was no significant difference in the rate of HSF among the three groups. Fig 1B shows the percentage of participants who met FAT criteria for each number of items. The percentage of participants who met FAT criteria in two items was significantly higher in the group of rhythmic gymnasts than in the other two groups (rhythmic gymnasts [43.6%] vs. volleyball players [13.0%] vs. non-athletes [16.8%], *p*<0.05), and the percentage of participants who did not meet any FAT criteria was significantly lower in the group of rhythmic gymnasts than in the other two groups (rhythmic gymnasts [15.4%] vs. volleyball players [41.3%] vs. non-athletes [43.0%], *p*<0.05).

**Fig 1.**
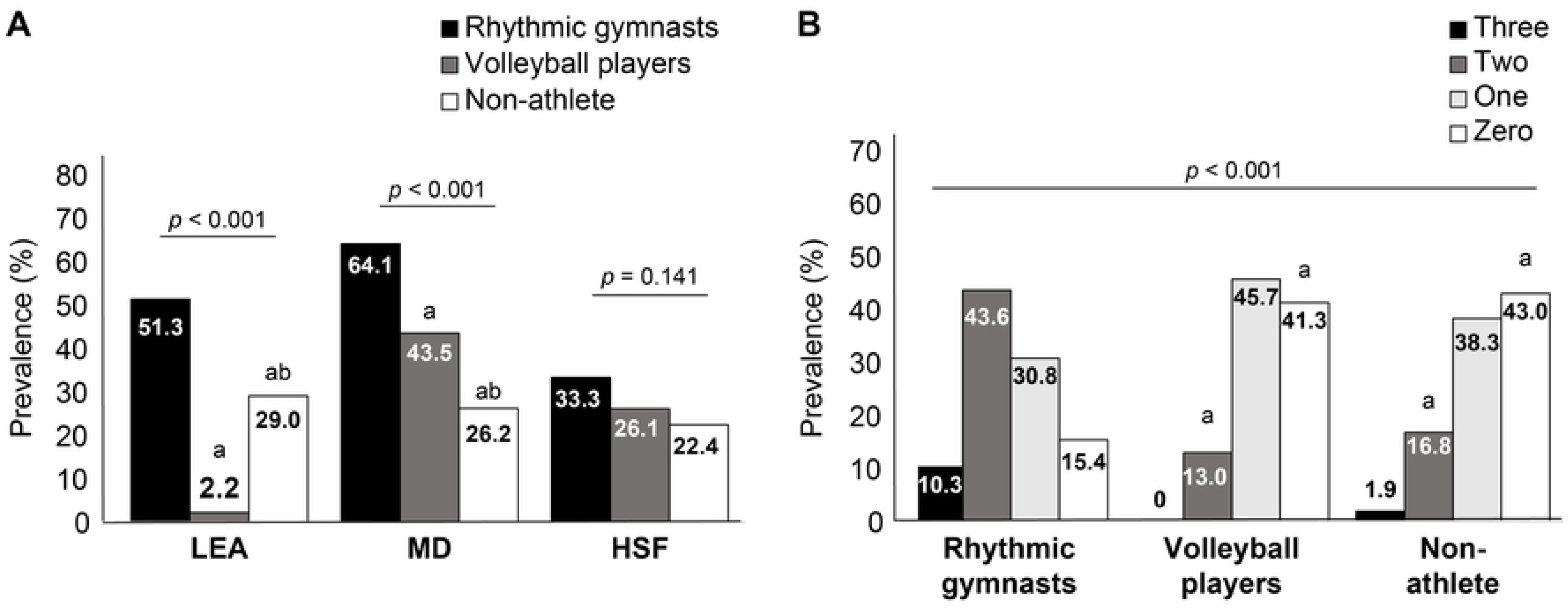
The prevalence of athletes with FAT and the number of those at FAT-related risk. A. LEA <85% of standard body weight (BMI=22 kg/m^2^); MD, including amenorrhea and menstrual irregularities; and HSF ^a^*p*<0.05 rhythmic gymnasts; ^b^*p*<0.05 vs. volleyball, players, using a χ^2^ test with Bonferroni correction in each group B. The prevalence of each number at risk of the FAT ^a^*p*<0.05 vs rhythmic gymnasts, using a χ2 test with Bonferroni correction in each group Abbreviations: BMI, body mass index; FAT, female athlete triad; HSF, history of stress fracture; LEA, low energy availability; MD, menstrual dysfunction.

### Intergroup comparisons for BI and eating behaviors

Table 2 shows a comparison between BI and eating behaviors across the three groups. Significant differences were observed in I-BI scores, with the rhythmic gymnasts reporting a significantly lower score than those in the other two groups (rhythmic gymnasts [2.8±0.9] vs. volleyball players [4.4±0.9] vs. non-athletes [3.8±1.2], *p<*0.001). Additionally, the BI distortion score was significantly higher in the rhythmic gymnasts compared with the volleyball players (rhythmic gymnasts [0.7±1.2] vs. volleyball players [-1.0±1.1] vs. non-athletes [0.2±1.4], *p*<0.001), indicating that the rhythmic gymnasts overestimated their body shape.

**Table 2.**
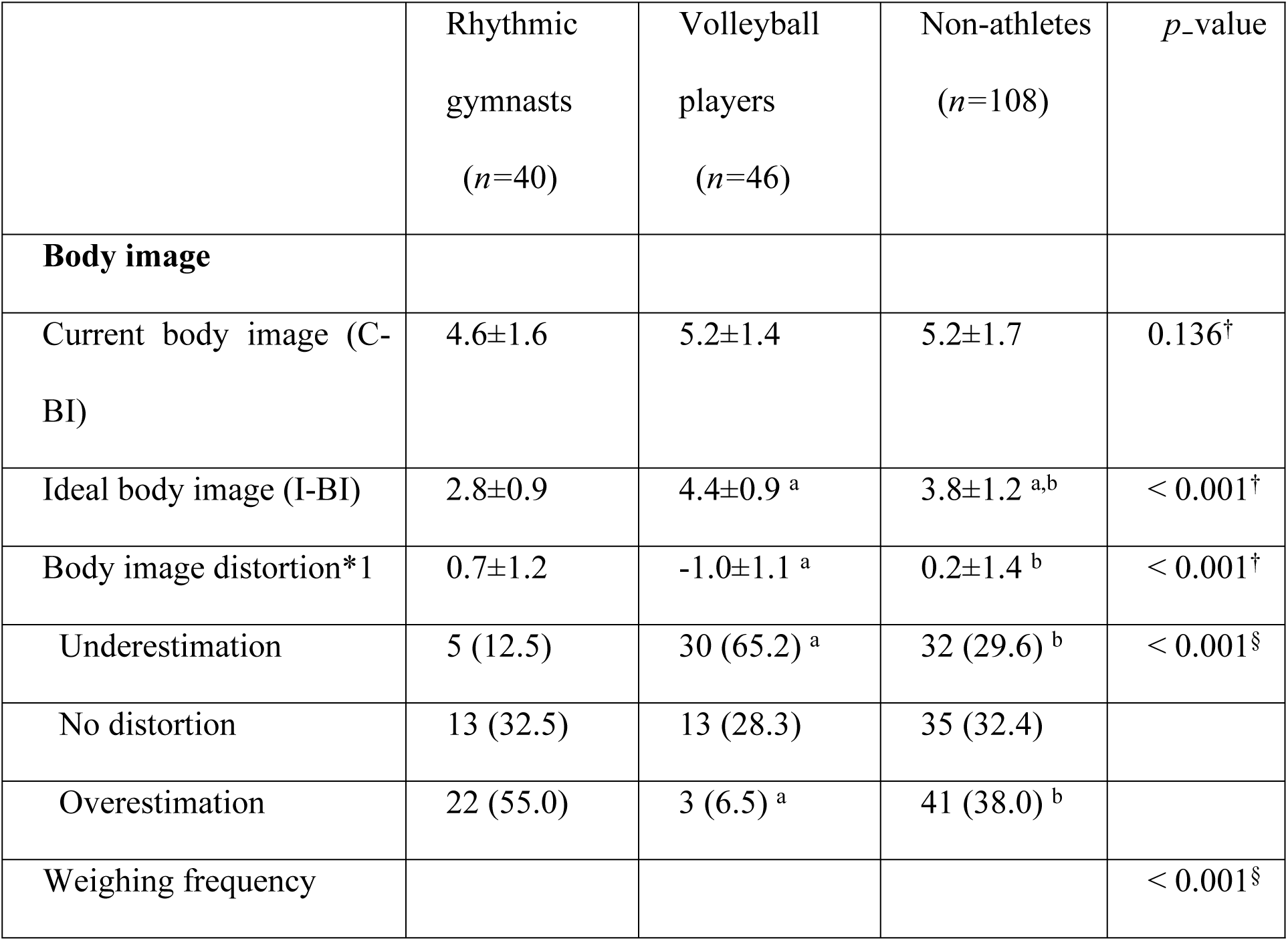
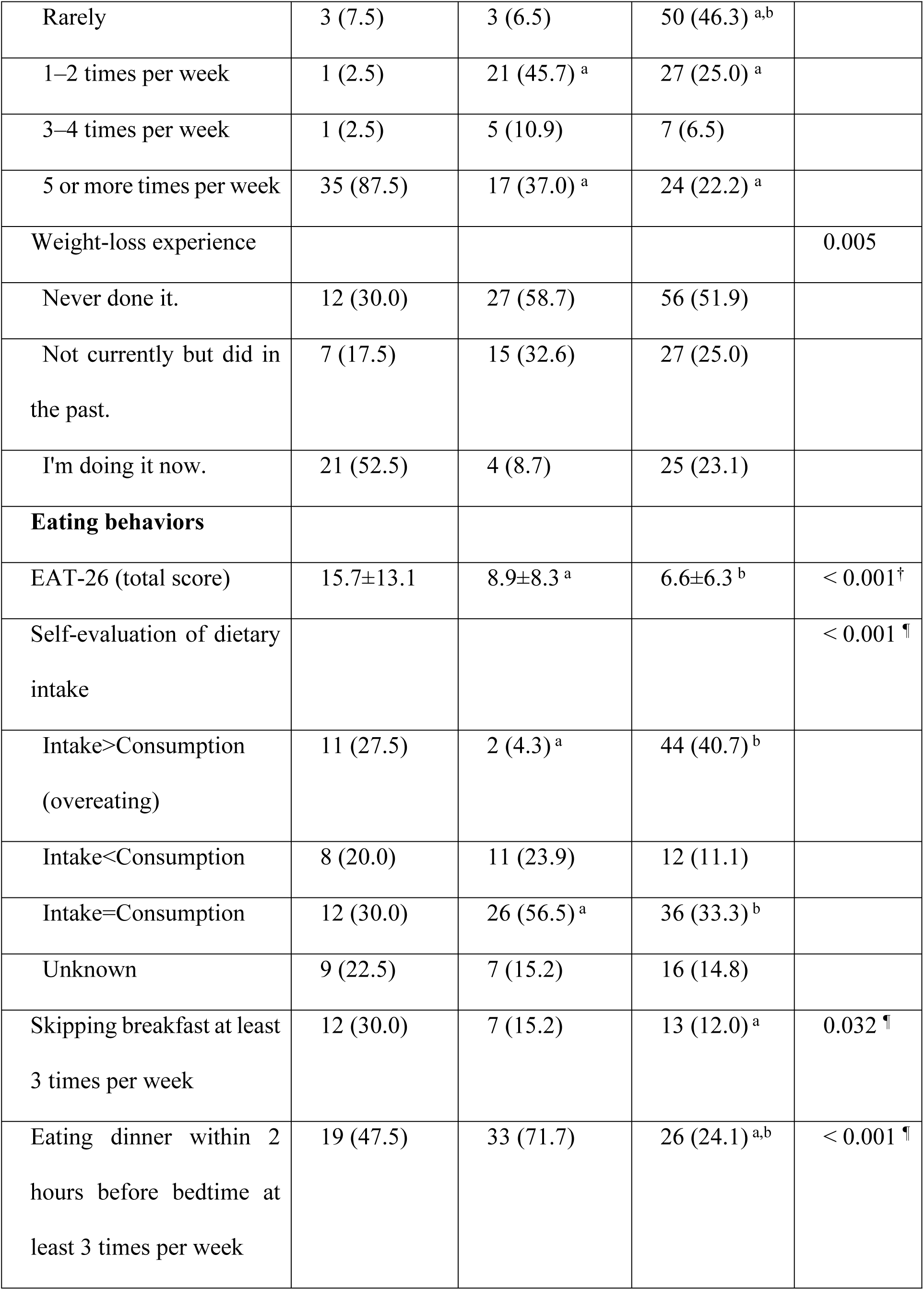
Body image and eating behaviors in rhythmic gymnasts, volleyball players, and non-athletes.

The proportion of participants who overestimated their body shape was significantly higher in the rhythmic gymnasts (55.0%) than in the volleyball players (6.5%) (*p*<0.05) and non-athletes (38.0%) (*p*<0.05). The proportion of individuals who measured their weight ≥5 times a week was significantly higher in the rhythmic gymnasts (87.5%) than in the volleyball players (37.0%) and the non-athletes (22.2%) (*p*<0.001). Furthermore, the rhythmic gymnasts had a significantly higher percentage of individuals who had experienced weight loss (52.5%) compared with the volleyball players (8.7%) and the non-athletes (23.1%) (*p*=0.005).

In terms of eating behaviors, the rhythmic gymnasts had a significantly higher EAT-26 score than those in the other groups (rhythmic gymnasts [15.7±13.1] vs. volleyball players [8.9±8.3] vs. non-athletes [6.6±6.3], *p*<0.001). The percentage of participants who self-evaluated their food intake as exceeding their consumption (overeating) was significantly higher in the rhythmic gymnasts (27.5%) compared with the volleyball players (4.3%) (*p*<0.001). The percentage of participants who skipped breakfast ≥3 times a week was also significantly higher in the rhythmic gymnasts (30.0%) compared with the non-athletes (12.0%) (*p=*0.032). The percentage of participants who ate dinner 2 h before bedtime was significantly higher in both the rhythmic gymnasts (47.5%) and the volleyball players (71.7%) compared with the non-athletes (24.1%) (*p*<0.001). The rhythmic gymnasts consumed rice less frequently than those in the other two groups, but this difference did not reach statistical significance (*p*=0.058).

### FAT-related factors

Table 3 presents the results of the multivariate logistic regression analysis in relation to each FAT component as a dependent variable. In a comparison between rhythmic gymnasts and volleyball players (*n*=86), LEA was independently associated with being a rhythmic gymnast and with overestimation of body shape. MD was independently associated with high EAT-26 scores. No significant association was found for HSF. In a comparison between rhythmic gymnasts and non-athletes (*n*=148), LEA was independently associated with being a rhythmic gymnast and with overestimation of body shape. MD was independently associated with being a rhythmic gymnast and with high EAT-26 scores. HSF was independently associated with high EAT-26 scores.

**Table 3.** Risk factors associated with female athlete triad (FAT).

|  | LEA |  | MD |  | HSF |  |
| --- | --- | --- | --- | --- | --- | --- |
|  | OR<br>(95%CI) | *p*-value | OR<br>(95%CI) | *p*-value | OR<br>(95%CI) | *p*-value |
| Rhythmic gymnasts<br>and volleyball players<br>(*n*=86)<sup>†</sup> | (*n*=86) |  | (*n*=84) |  | (*n*=86) |  |
| Rhythmic gymnasts<br>(vs. volleyball players) | **174.41**<br>**(6.38-927.83)** | **0.003** | 1.09<br>(0.31-3.82) | 0.894 | 0.98<br>(0.27-3.57) | 0.981 |
| Body image distortion<sup>Φ</sup> | **4.34**<br>**(1.60-11.80)** | **0.004** | 1.36<br>(0.86-2.14) | 0.190 | 1.00<br>(0.65-1.53) | 0.997 |
| EAT-26 (total score)<sup>Φ</sup> | 0.93<br>(0.92-1.10) | 0.931 | **1.07**<br>**(1.01-1.14)** | **0.024** | 1.04<br>(0.99-1.09) | 0.087 |
| Rhythmic gymnasts | (*n*=148) |  | (*n*=146) |  | (*n*=148) |  |
| and non-Athlete (*n*=148)<sup>‡</sup> |  |  |  |  |  |  |
| Rhythmic gymnasts<br>(vs. non-athlete) | **3.95**<br>**(1.02-15.36)** | **0.047** | **3.18**<br>**(1.36-7.56)** | **0.009** | 1.21<br>(0.46-3.17) | 0.698 |
| Body image distortion<sup>Φ</sup> | **7.45**<br>**(3.63-15.30)** | **<0.001** | 1.24<br>(0.94-1.66) | 0.134 | 0.81<br>(0.60-1.10) | 0.179 |
| EAT-26 (total score)<sup>Φ</sup> | 1.05<br>(0.97-1.14) | 0.210 | **1.06**<br>**(1.01-1.11)** | **0.024** | **1.04**<br>**(1.00-1.09)** | **0.047** |

<sup>†</sup> Adjusted for age and athletic career, <sup>‡</sup> Adjusted for age, <sup>Φ</sup> Data continuous
LEA, low energy availability; MD, menstrual dysfunction; HSF, history of stress fractures; OR, odds ratio; 95% CI,
EAT; Eating Attitude Test

## Discussion

The results of this study showed that female rhythmic gymnasts were thinner than both female volleyball players and non-athletes and exhibited a higher prevalence of each FAT component-related sign, particularly LEA and MD. Rhythmic gymnasts, similar to non-athletes, reported a greater overestimation of their own body shape compared with female volleyball players. Furthermore, logistic regression analysis revealed that being a rhythmic gymnast, overestimating body shape, and engaging in eating disorder-related eating behaviors were independently associated with LEA or MD. We identified a significantly higher prevalence of each FAT component, especially LEA and MD, among the rhythmic gymnasts. Approximately 50% of the rhythmic gymnasts met ≥2 FAT criteria. This finding is consistent with findings reported elsewhere that indicate a higher prevalence of LEA (BMI <17.5 kg/m²) in Japanese university athletes of aesthetic sports (7.2%), which exceeds that of other sports [14] and a greater proportion meeting multiple FAT criteria compared with athletes of other sports [9]. The rigorous training and aesthetic demands of rhythmic gymnastics lead to increased energy expenditure, while the need to maintain a specific BI often leads to insufficient energy intake, thereby resulting in LEA.

The association between LEA and MD has been previously reported [5,17,18]. Irregularities and abnormalities in the menstrual cycle typically occur when energy expenditure from exercise exceeds energy intake over a certain period (relatively long), which is consistent with our findings. However, no significant difference in the prevalence of HSF was observed. Despite the high-impact nature of rhythmic gymnastics, which places substantial stress on joints and bones, it is possible that athletes of such sports have higher bone density compared with those in non-impact sports such as swimming or non-athletes [19,20]. Additionally, the effects on bone may manifest later than concerns such as menstrual abnormalities [21]. Measuring bone density with high-precision equipment such as dual-energy x-ray absorptiometry may be necessary to detect these issues. Furthermore, HSF is influenced by factors such as calcium and vitamin D intake [22] and exercise intensity [9]. suggesting that further research is needed to account for these variables. With regard to BI, >50% of the rhythmic gymnasts in this study overestimated their body shape, which was a significantly higher proportion among volleyball players. A previous study reported that Japanese non-athlete females also tend to overestimate their body shape, particularly at a younger age, and that a desire to lose weight among thin women (BMI <18.5 kg/m²) is associated with distorted BI perceptions [7]. Our study findings suggest that, similar to non-athlete women, rhythmic gymnasts’ overestimation of their body shape may exacerbate their desire to lose weight. Additionally, rhythmic gymnasts had a lower BMI compared with volleyball players and non-athletes, and their ideal body shape was notably slender. They also reported more frequent weight measurement and the highest proportion of individuals currently dieting, with >50% engaged in weight loss efforts. These results align with the findings of Bruin et al. [23] that, despite having a lower BMI than the general population, female athletes of aesthetic sports often have an increased desire to lose weight and engage in dieting more frequently.

Distorted BI, body dissatisfaction, and weight concerns are well-documented risk factors for eating disorder. BI disturbances have been linked to eating disorder and weight loss behaviors in athletes [24,25] and, among athletes of aesthetic spots, the desire to present a “perfect” body shape is known to exacerbate tendencies toward eating disorder [26]. In this study, rhythmic gymnasts exhibited higher EAT-26 scores, eating disorder, and reported a higher frequency of skipping breakfast compared to the other two groups, indicating disordered eating patterns. These findings suggest that the nature of rhythmic gymnastics contributes to a belief among athletes that weight loss is necessary for performance enhancement. This distorted BI perception may further intensify their weight loss efforts, leading to inadequate eating behaviors such as food restriction.

Factors associated with the prevalence of each FAT components include participation in rhythmic gymnasts, distorted BI, and eating behaviors linked to eating disorder. Previous studies have identified inappropriate eating behaviors as risk factors for FAT, noting that such behaviors can alter hormone levels, affect the reproductive system, and ultimately disrupt the menstrual cycle, which may negatively affect bone health if not corrected [27]. This study similarly highlights that distorted BI contributes to LEA, a primary factor in FAT. The results imply that rhythmic gymnasts’ desire for a lean body to enhance competitive performance, coupled with an overestimation of their body shape, may precipitate the development of FAT. These findings underscore the need for targeted support for young female athletes in Japan, particularly rhythmic gymnasts, to promote healthy BI and eating behaviors to prevent FAT.

This study had several limitations. We relied on an online survey in which participants reported their menstrual status and HSF without formal medical diagnosis. A detailed dietary survey was not conducted, meaning that energy availability was not quantitatively assessed. Despite these limitations, to the best of our knowledge, this study is the first to examine the prevalence of FAT and its association with BI among young female athletes involved in rhythmic gymnastics and volleyball and among non-athletes.

### Conclusion

The prevalence of FAT (at least one or more components) was approximately 1.5 times higher among rhythmic gymnasts than among volleyball players and non-athletes. Logistic regression analysis indicated that FAT-associated factors include participation in rhythmic gymnasts, distorted BI, and eating behaviors linked to eating disorder. These findings suggest that future support for appropriate body image and eating behaviors among Japanese rhythmic gymnasts is required.

## Data Availability

The datasets generated and analysed during the current study are not publicly available as the University of Hyogo's research ethics committee requires to protect personal data. But the data are available from the corresponding author on reasonable request.

## Acknowledgments

None.

